# SECOND ALLOGENEIC STEM CELL TRANSPLANTATION: A MORTALITY ANALYSIS

**DOI:** 10.1101/2020.07.21.20153684

**Authors:** Mustafa Alani, Jean Henri Bourhis

## Abstract

Second allogeneic stem cell transplantation was realized in 48 patients with myeloid and lymphoid neoplasms at Gustave Roussy institute since 1987. Overall survival rate was about 30 % with better outcome in acute myeloid leukemia cases. Non-relapse related mortality is overwhelming, especially in myelodysplasia patients and despite the fact that complete remission was obtained in their majority. Graft versus Host disease is very common after second transplantation with many grade III – IV cases and one death from severe pulmonary GvHD lesions. Reduced intensity conditioning is certainly less toxic and together with optimal GvHD and infectious disease management, Second SCT may be a reasonable therapeutic option and the only curative treatment for many hematological malignancies.

## Introduction

Allogeneic Stem Cell Transplantation is the only curative treatment in many hematological disorders. Yet, mortality rate is non-negligible.

In recent years, many retrospective studies showed interesting findings with prolonged remission and very manageable toxicity in second transplanted patients ^(2)^.

Second allogeneic SCT has now become a reasonable therapeutic option. More bridging strategies are available. The age limit is expanding, thanks to improving supportive care. In myeloid malignancies, getting complete remission is more possible than ever. Making the most of it, therefore, is tempting.

The following is a retrospective single-center cohort study on 48 second transplanted patients in Gustave Roussy Institute, from 1987 to 2019. Focusing on remission rate, post-transplant events, Graft versus Host disease, causes of mortality and non-relapse mortality. Trying to improvise some aiding strategies. We are considering mainly myeloid malignancy patients, on whom; the data is relatively larger and more informative.

## Background and State of the Art – Literature review

### Patients and Methods

The study materiel consists of 48 second transplantations, done during 32 years for patients with relapsed myeloid and lymphoid malignancies, at Gustave Roussy Cancer Center (1987 – 2019).

Overall survival was the first finding, together with final complete remission rate, remission before transplantation, bridging treatments, duration of remission and causes of death.

Secondarily, we looked for patient ages, graft sources, transplantation years, conditioning protocols, GvH disease and viral reactivations.

Reported complete remission in acute leukemia cases is at least cytological. This implies myelodysplastic syndromes. In chronic myeloid leukemia, chronic phase is also considered a remission. Complete remission in myeloma patients is biological, hematological and radiological. Complete remission in lymphoma patients is morphological. All other disease states were considered advanced disease.

Conditioning protocols and GvHD grading were defined according to EBMT standards.

## Results

Within 48 patients, only 31.2 % (15 patients) are still alive. 70.8 % (34 patients) had complete remission after second transplantation. 2 % had partial response (one multiple myeloma patient), 2 % had progressive disease (one diffuse large B cell lymphoma patient), 18.7 % (9 patients) had no response assessment in the short period before death. Finally, 3 graft failure / no-engraftment cases were observed (table 1) (figure 1, 5 and 6).

**Table 1.**
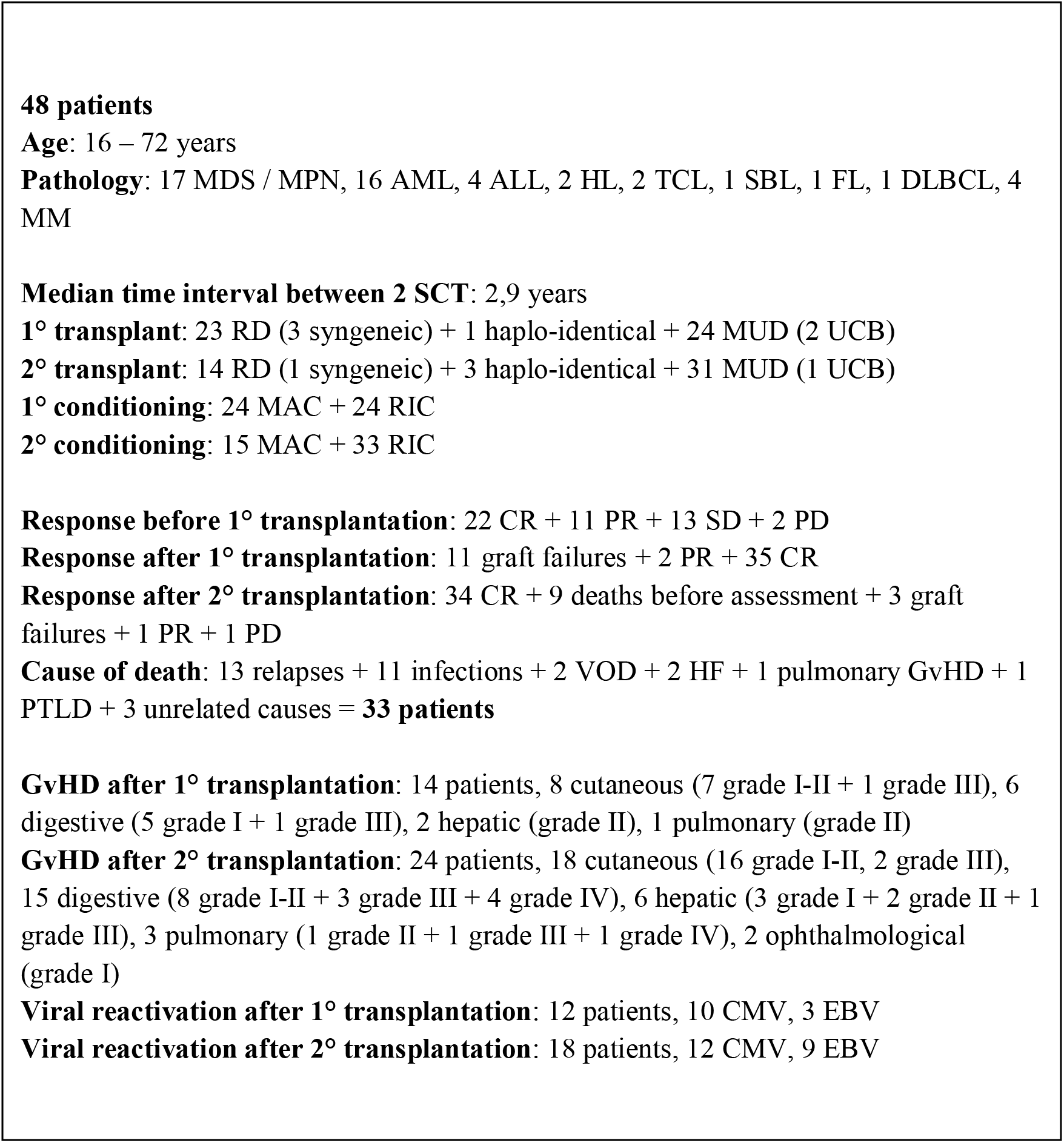
All patients’ transplantation main events and characteristics.

**Figure 1.**
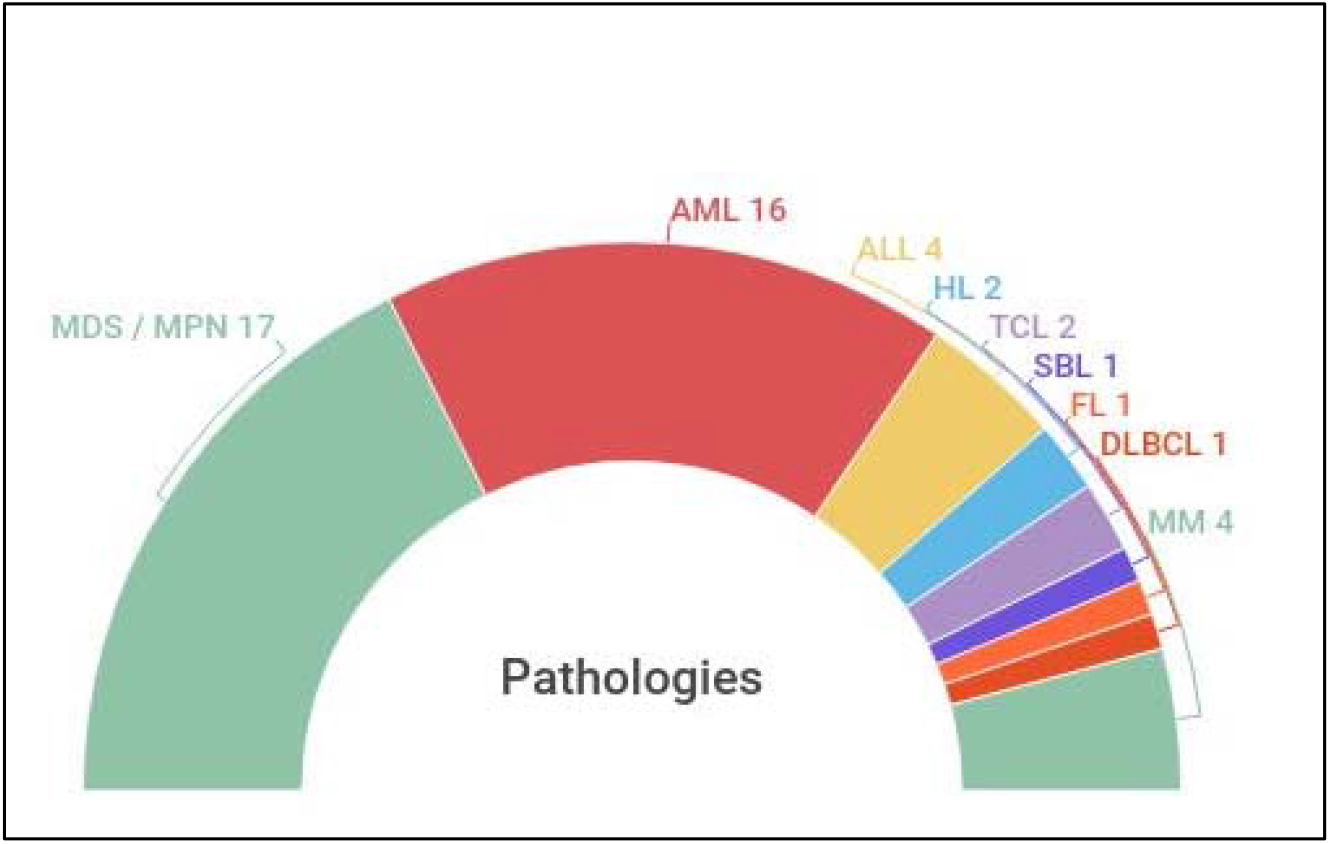
Pathologies in second transplanted patients

24 patients (50 %) did not lose their first graft and were secondarily transplanted due to relapse / progressive disease.

12 patients lost their first graft after D100, six of them had salvage treatment and complete remission was obtained in two.

10 patients lost their first graft before D100, one of them had salvage treatment (Azacitidine, AML patient) leading to complete remission before second transplantation.

Finally, two patients (AML patients) were rescued by Donor Lymphocyte Infusions with chemotherapy and did not lose their first graft.

Patient age upper limit was 72 years old (figure 2).

**Figure 2.**
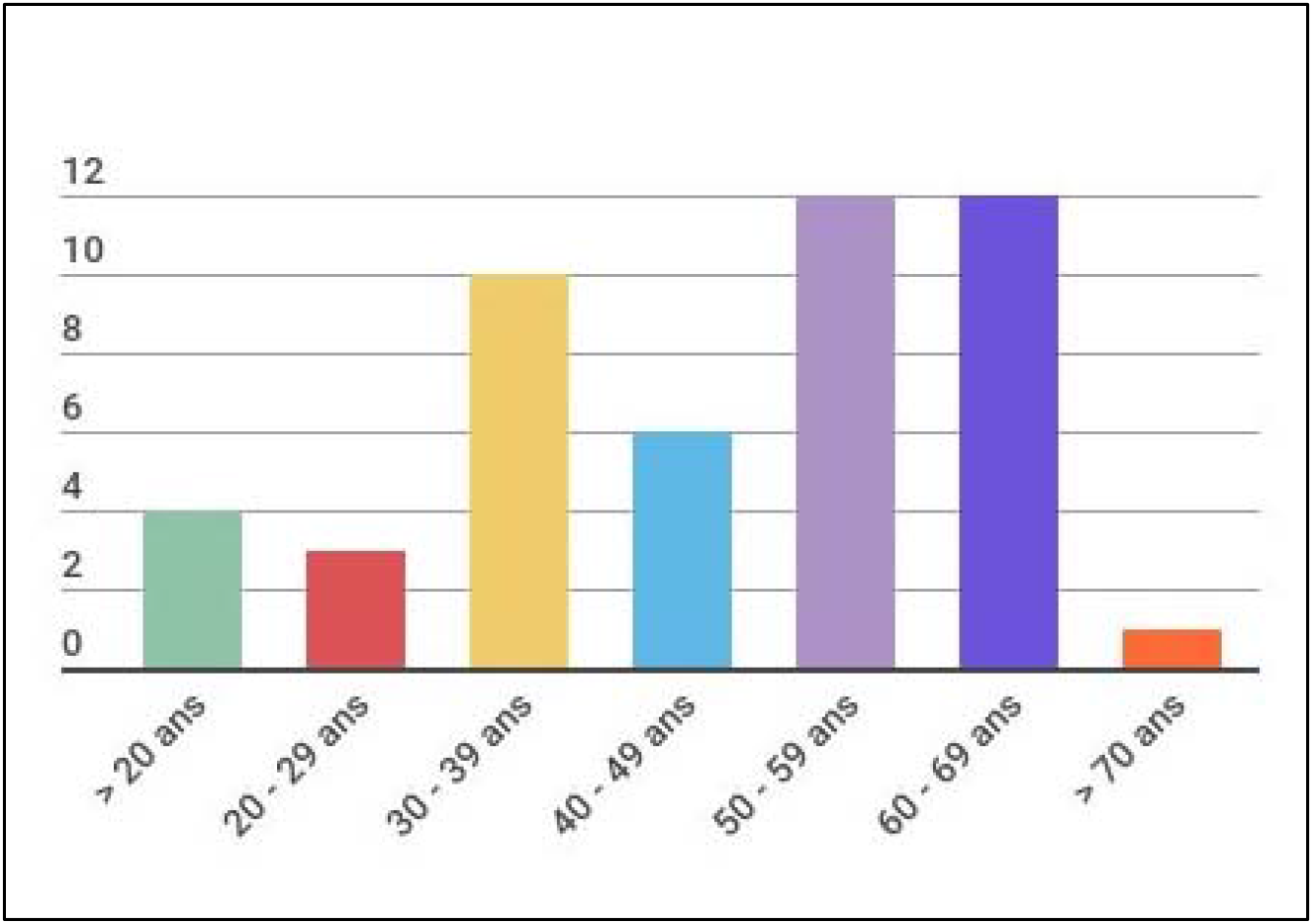
Patient ages

Median time interval between two transplantations is 2.9 years. A little shorter in alive patients (2.4 years).

5.4 years is the median follow-up time for alive patients after second transplantation. Cause of death in 27 % (13 patients) is progressive malignancy and relapse. 11 patients (22.9 %) are dead from bacterial infections during or shortly after transplantation. 2 patients died from veno-occlusive disease. 2 from sever heart failure. One death from sever pulmonary GvH disease. One from a rapidly progressive Post Transplantation Lymphoproliferative Disorder and 3 deaths from unrelated causes (accident, second neoplasia…) (figure 4). Transplantation years are illustrated in (figure 3), showing significant correlation with the rate of survival (fewer deaths with advanced supportive care).

**Figure 3.**
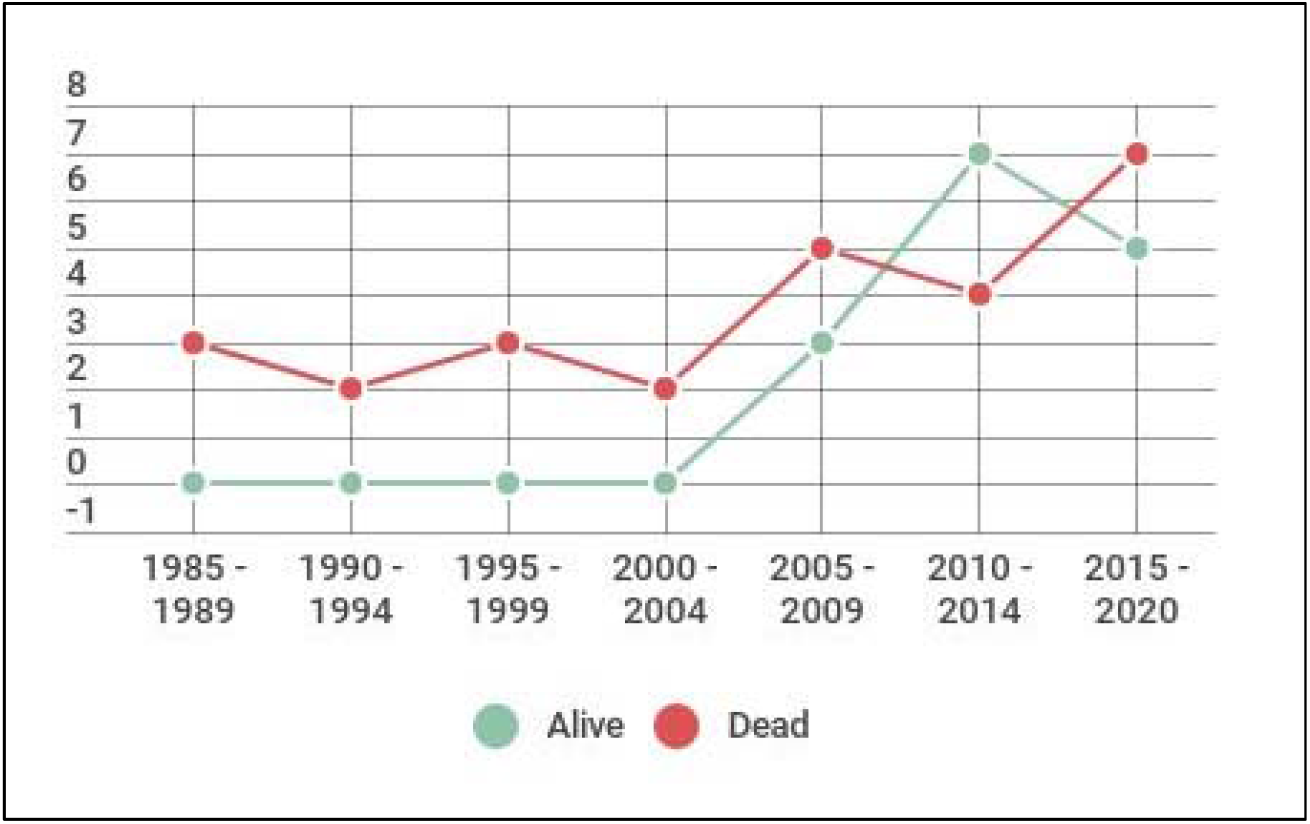
Year of transplantation and OS in second transplanted patients

**Figure 4.**
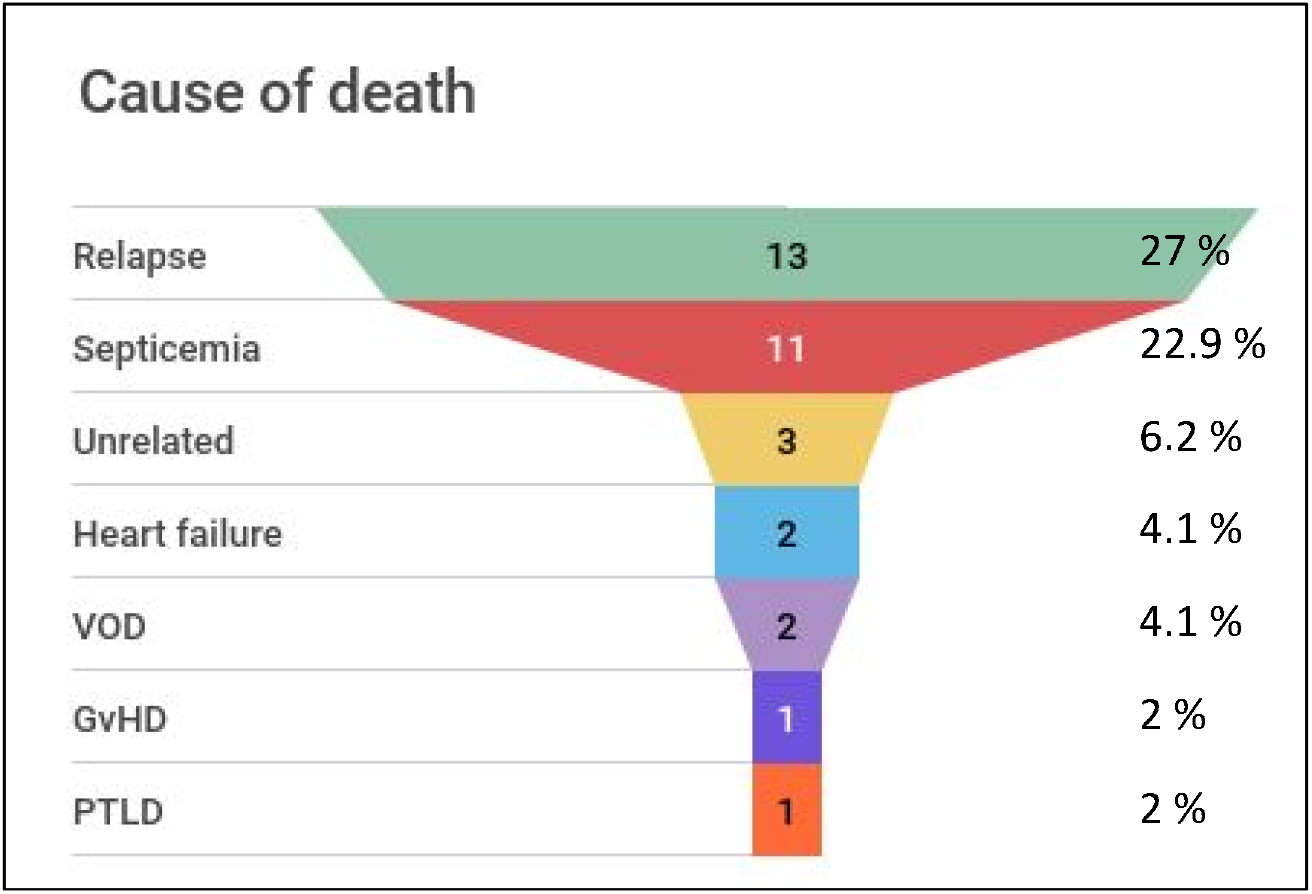
Death causes in second transplanted patients

GvHD and viral reactivation cases are illustrated in (table 1 and 2). Second SCT nearly doubles the cases of GvHD and EBV/CMV reactivation. This has no impact on remission rate or final outcome whatsoever.

**Table 2.**
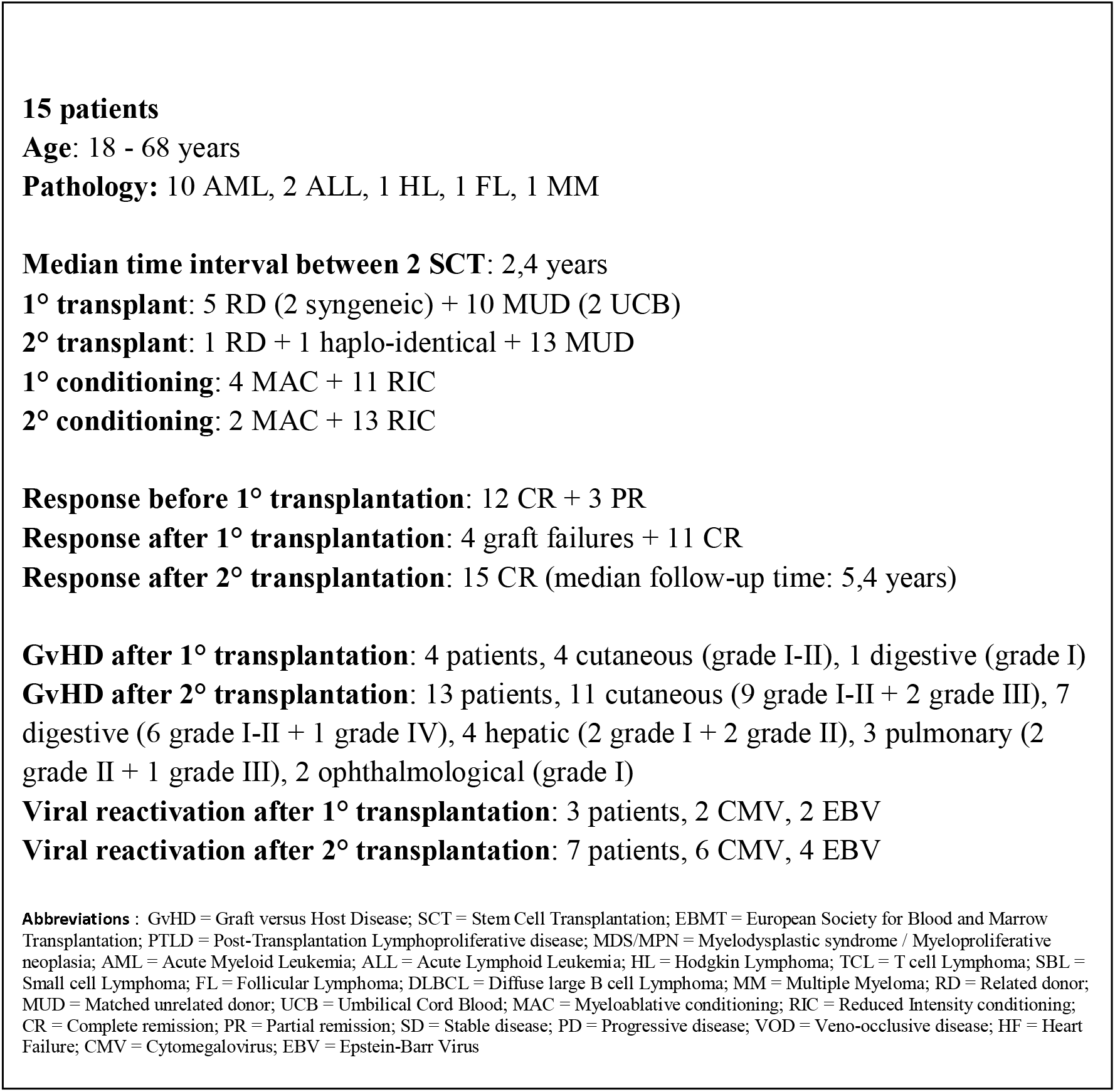
Alive patients’ transplantation main events and characteristics

Conditioning protocols are illustrated in (table 1 and 2).

Alive patients received mostly RIC regimens (4 MAC + 11 RIC in first transplantation, 2 MAC + 13 RIC in second transplantation).

Graft sources are illustrated in (table 1 and 2), showing larger number of unrelated donors without remarkable impact on the occurrence of GvHD.

We had 17 MDS/MPN cases and 16 AML cases. Complete remission was observed in 1 MDS/MPN case and 15 AML cases (93.7 %) after initial treatment before first transplantation (figure 7).

**Figure 5.**
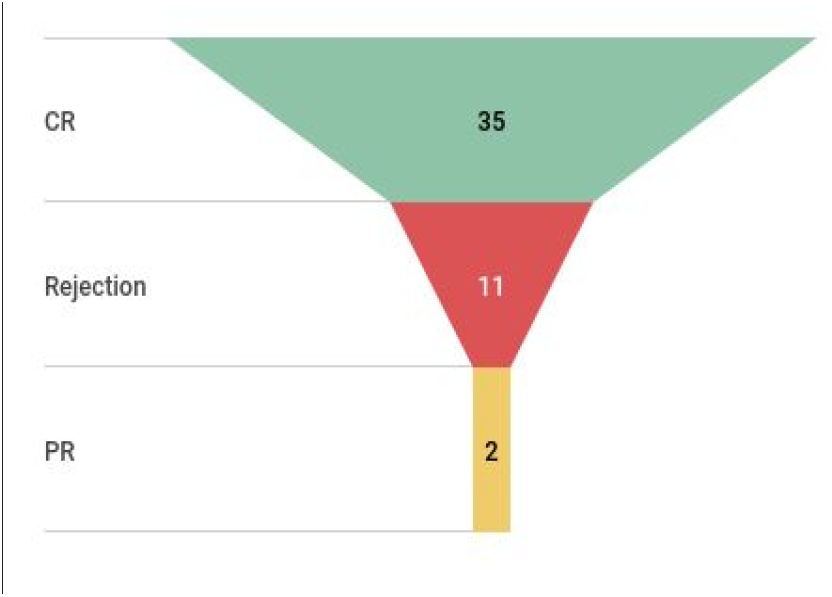
Response after 1^st^ transplantation.

**Figure 6.**
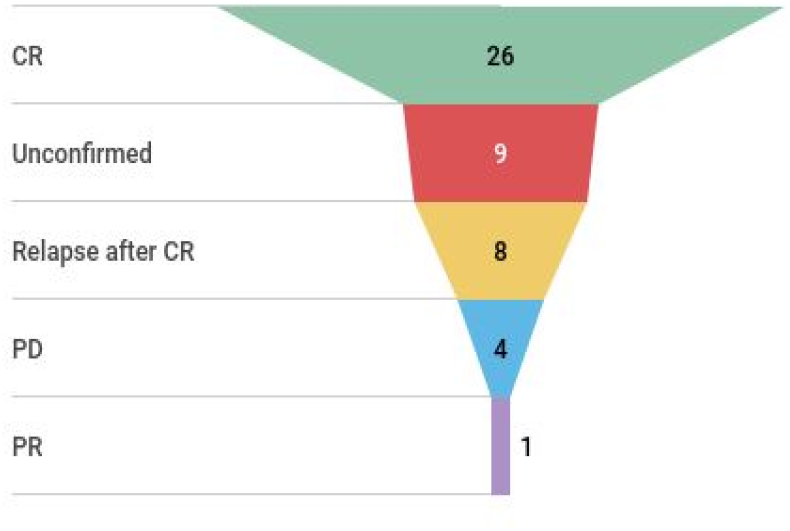
Response after 2^nd^ transplantation

**Figure 7.**
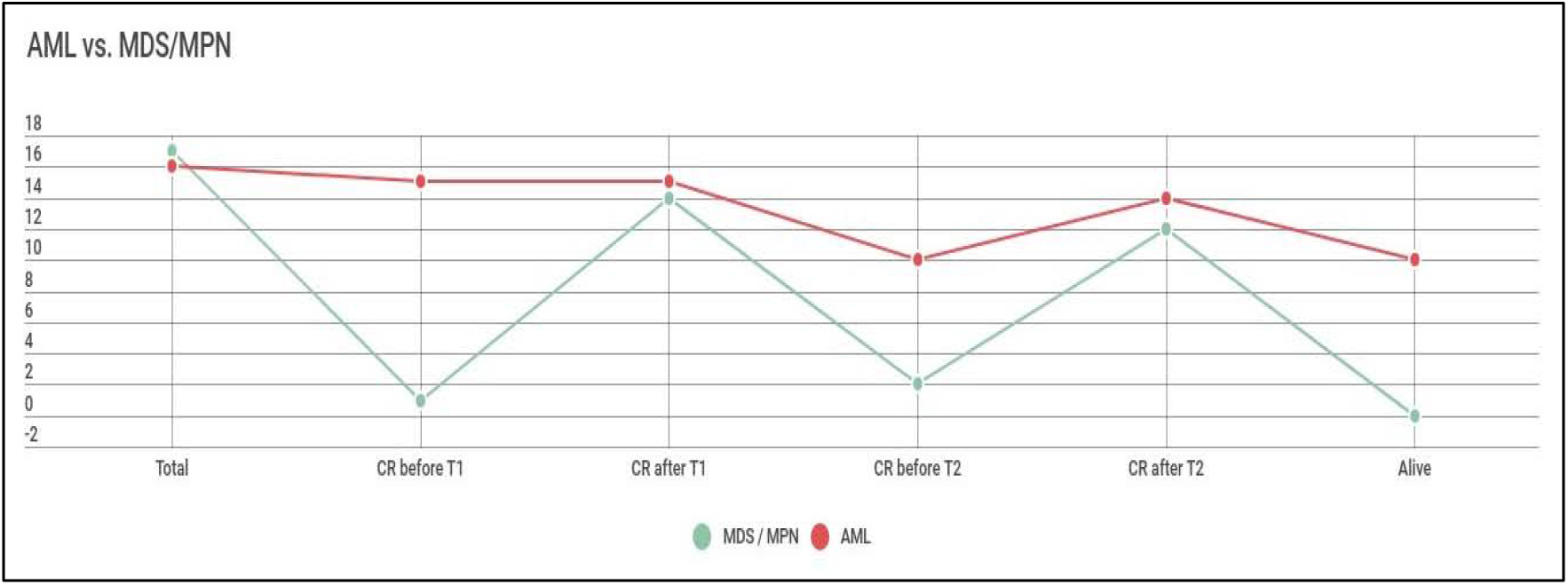
Response and Mortality in AML and MDS/MPN patients, in relation with SCT

First line treatment in MDS/MPN patients included: Hydroxyurea, Interferon-alpha, Tyrosine kinase inhibitors, Low-Dose Cytarabine and Azacitidine.

Treatments in AML patients were mainly the classical 7 + 3 induction chemotherapy (81.2 %). One associated with Gemtuzumab ozogamicine. One patient received the VANDA regimen and two patients had the CLARA regimen.

Salvage regimens included mainly Azacitidine, Hydroxyurea and 7 + 3 chemotherapy in MDS/MPN patients.

AML patients were treated by CLARA regimen, FLAG regimen, VALOR regimen, Low-Dose Cytarabine with or without Gemtuzumab ozogamicine, Azacitidine and Decitabine. Complete remission was obtained in 15 AML cases (93.7 %) and 14 MDS/MPN cases (82.3 %) after first transplantation.

Bridging treatment allowed second complete remission in 10 AML and 2 MDS/MPN cases. Second transplantation allowed complete remission in 14 AML (87.5 %) and 12 MDS/MPN cases (70.5 %) (figure 7).

However, all 12 MDS/MPN patients died from non-relapse related mortality (6 from severe infections, 1 VOD, 1 PTLD, 1 Heart failure, 1 Pulmonary GvHD and 2 unrelated causes). 3 AML patients died from late relapse (6 months, 10 months and 35 months after second SCT).

## Discussion

Allogeneic SCT is still the only curative treatment for most myeloid malignancies ^(3)^. For first transplantation, the decision is relatively simple. Whereas, second transplantation requires more attention. From the data above and other related studies, the age is irrelevant ^(4)^.

In our single center retrospective study, there was no relation between overall survival and time interval between first and second transplantation. Interestingly, this later was a bit shorter in alive patients ^(2)^.

Second transplantation nearly doubles the risk of GvHD ^(5)^. 50 % of patients in our data had a significant GvHD, seven of which (30 %) had a Grade III – IV disease (table 1 and 2). Although, death due to severe GvHD was observed in only one patient (a MDS patient). Optimal GvHD management remains therefore, very essential.

Better outcome was observed in AML patients ^(6)^, regarding complete remission rate and overall survival after second transplantation.

MDS/MPN patients had interestingly high complete remission rate after second transplantation. They had also the most significant non-relapse related mortality. However, improvements in infectious disease management and supportive care result in better outcome in the last two decades.

Reduced intensity conditioning ^(7)^ is largely used for about 20 years now and NRM rates are lower, certainly in patients with comorbidities. Regarding hematological outcome however, beside its higher toxicity, no better result was obtained from myeloablative protocols, at least in present data ^(8)^.

Graft sources are now more available than ever, unrelated and especially haploidentical stem cell donors ^(9)^. This led to more organized transplantation programs, accelerate procedures and certainly improve outcome in patients with no related donor available.

## Conclusion

Current data shows a non-negligible good outcome in second transplanted myeloid malignancy patients, who despite new treatments today, still have a high risk of relapse without transplantation ^(3)^.

GvHD is very common in second transplanted patients and optimal management is crucial. Reduced intensity conditioning has to be considered in second transplantation, for a lower rate of NRM.

## Data Availability

research data is available in Gustave Roussy registry

## Abbreviations

GvHD: Graft versus Host Disease
SCT: Stem Cell Transplantation
EBMT: European Society for Blood and Marrow Transplantation
PTLD: Post-Transplantation Lymphoproliferative disease
MDS/MPN: Myelodysplastic syndrome / Myeloproliferative neoplasia
AML: Acute Myeloid Leukemia
ALL: Acute Lymphoid Leukemia
HL: Hodgkin Lymphoma
TCL: T cell Lymphoma
SBL: Small cell Lymphoma
FL: Follicular Lymphoma
DLBCL: Diffuse large B cell Lymphoma
MM: Multiple Myeloma
RD: Related donor
MUD: Matched unrelated donor
UCB: Umbilical Cord Blood
MAC: Myeloablative conditioning
RIC: Reduced Intensity conditioning
CR: Complete remission
PR: Partial remission
SD: Stable disease
PD: Progressive disease
VOD: Veno-occlusive disease
HF: Heart Failure
CMV: Cytomegalovirus
EBV: Epstein-Barr Virus

## References

1. Duus JE et al. Second allografts for relapsed hematologic malignancies: feasibility of using a different donor. Bone Marrow Transplantation 2005; 35: 261–264.

2. Ruutu T et al. Second allogeneic transplantation for relapse of malignant disease: retrospective analysis of outcome and predictive factors by the EBMT. BMT 2015; 50: 1542–1550.

3. Mielcarek M et al. Outcomes among patients with recurrent high-risk hematologic malignancies after allogeneic hematopoietic cell transplantation. Biology of Blood and Marrow Transplantation 2007;13(10):1160–8.

4. Sorror ML et al. Prospective Validation of the Predictive Power of the Hematopoietic Cell Transplantation Comorbidity Index: A Center for International Blood and Marrow Transplant Research Study. Biology of Blood and Marrow Transplant 2015; 21(8):1479–87.

5. Martin S et al. Bone Marrow GvHD after Allogenic Hematopoietic Stem Cell Transplantation. Frontiers in Immunology 2016; 7: 118.

6. Schneidawind C et al. Second allogeneic hematopoietic cell transplantation enables long-term diseasefree survival in relapsed acute leukemia. Annals of Hematology 2018; 97(12): 2491–2500.

7. Erden A et al. A Review of Myeloablative vs Reduced Intensity/Non-Myeloablative Regimens in Allogeneic Hematopoietic Stem Cell Transplantations. Balkan Medical Journal 2017; 34(1): 1–9.

8. Shaw BE et al. Outcome of second allogeneic transplants using reduced-intensity conditioning following relapse of hematological malignancy after an initial allogeneic transplant. Bone Marrow Transplantation 2008; 42(12): 783–9.

9. Thomas R. Haploidentical Stem Cell Transplantation: The Always Present but Overlooked Donor. Hematology ASH Education Program 2005; 390–5.

